# Fine-scale spatial mapping of Anaemia in two sentinel communities within a urogenital schistosomiasis-endemic area in Malawi

**DOI:** 10.64898/2026.08.19.26360771

**Authors:** Farah Khalid, Seke A. Kayuni, Peter Makaula, Janelisa Musaya, Sarah Rollason, J. Russell Stothard, Alexandra Brown, Emanuele Giorgi

**Author notes:** Corresponding author: (FK).

## Abstract

While a causal link between urogenital schistosomiasis (*Schistosoma haematobium*) and anaemia is well established, quantitative associations between infection intensity and haemoglobin levels across endemic communities, at fine spatial scales, remain insufficiently characterized. As part of the broader Hybridisation in Urogenital Schistosomiasis (HUGS) investigation, we studied the micro-epidemiology of anaemia within two study communities in southern Malawi where *S. haematobium* remains endemic.

Urine samples were examined by microscopy to quantify *S. haematobium* infection intensity, categorised as low (0–9 eggs/10 mL), moderate (10–49 eggs/10 mL), and heavy (≥50 eggs/10 mL). Individual haemoglobin concentrations were measured using a HemoCue photometer in 1,149 participants from Samama village (Mangochi District) and 977 participants from Mthawira village (Nsanje District). Linear geostatistical models incorporating individual-level characteristics and spatial covariates were used to estimate anaemia prevalence at fine geographical scales.

Moderate anaemia (Hb 80–109 g/L) was most prevalent among children aged 6–12 years (45.30% in Samama and 39.54% in Mthawira), while severe anaemia was more frequent among adults aged ≥19 years in both villages. Increasing *S. haematobium* infection intensity was associated with lower haemoglobin levels, with individuals harbouring heavy infections being at greater risk of moderate-to-severe anaemia. Spatial modelling revealed longer-range spatial correlation in Nsanje (φ = 66.7 km) than in Mangochi (φ = 25.5 km), indicating more spatially persistent risk in Nsanje and more localized heterogeneity in Mangochi. Predicted anaemia prevalence among female children aged 6–12 years with moderate infection intensity ranged from 25–65% in Mangochi and 64–76% in Nsanje.

At the micro-epidemiological level, *S. haematobium* infection intensity was associated with the severity of anaemia, with substantial spatial heterogeneity within and between villages. Identification of high-risk clusters supports targeted interventions, including stepped-up preventive chemotherapy, improved water and sanitation, and iron supplementation.

## Introduction

Anaemia, characterised by a reduced number of red blood cells or low haemoglobin (Hb) levels, impairs the body’s ability to transport oxygen efficiently. Globally, it affects an estimated 1.92 billion people, with the highest burden in low- and middle-income countries, particularly in sub-Saharan Africa (SSA)[1]. Within SSA, anaemia is especially prevalent among women of reproductive age and children under five, with pooled prevalence estimates of approximately 38–48% and 55% respectively, with substantially higher rates in high-burden countries[1, 2]. As the aetiology of anaemia in SSA is typically multifactorial, major contributors in young children include malaria, poor diet, and helminth infections ([3–5]. Among helminth infections, schistosomiasis is particularly important because of its disproportionate contribution to anaemia burden in high-transmission settings, substantially exceeding the population-level impact of soil-transmitted helminths in comparable endemic communities[4, 5]. Schistosomiasis is a neglected tropical disease (NTD) that remains highly endemic in Malawi and across much of SSA, with over 240 million people infected globally and more than 90% of cases occurring in SSA [5,6]. In Malawi, *Schistosoma haematobium* and *Schistosoma mansoni* are the predominant species, each with distinct lifecycles and clinical outcomes (6,7). Transmission of urogenital schistosomiasis occurs primarily along the margins and shorelines of Lake Malawi and the Lower Shire River, mediated by infected Bulinus freshwater snails, while S. mansoni is transmitted by Biomphalaria pfeifferi, both of which are locally present [6].

Specifically, urogenital schistosomiasis (*S. haematobium*) is a major contributor to anaemia due to sustained blood loss (haematuria) and chronic inflammation[7, 8]. Heavy infection intensity has been shown to be strongly linked to severe anaemia; for example, a study in Zanzibar demonstrated this association among school-age children[5]. Two sentinel communities in Mangochi and Nsanje districts were selected for detailed study as part of the Hybridisation in Urogenital Schistosomiasis (HUGS) investigation. These districts represent contrasting ecological and transmission settings: Mangochi lies along the southern shores of Lake Malawi, while Nsanje is situated along the Lower Shire River, providing an opportunity to examine spatial heterogeneity in anaemia risk across two distinct urogenital schistosomiasis-endemic environments. These communities are the focus of a longitudinal epidemiological study conducted against the background of ongoing preventive chemotherapy with praziquantel, delivered through mass drug administration (MDA) primarily targeting school-age children, highlighting the need for village-level mapping to guide and optimise intervention strategies [9–11].

Anaemia risk in schistosomiasis-endemic settings is influenced by spatially structured environmental exposure, particularly proximity to freshwater bodies where transmission occurs, alongside individual-level risk factors such as age and sex. The HUGS study provides a unique opportunity to investigate these patterns at fine spatial scales through georeferenced infection data and contemporaneous haemoglobin measurements, enabling spatial mapping of anaemia prevalence associated with urogenital schistosomiasis [9–11].

Despite biological and epidemiological evidence linking schistosomiasis with anaemia, fine-scale spatial risk mapping remains limited in Malawi, restricting effective targeting of interventions whose impact is already constrained by inflammation-driven iron sequestration (7, 10). Model-based geostatistics (MBG) offers a way to address this gap by integrating individual-level covariates with environmental data, allowing unmeasured spatial risk factors to be captured through Gaussian processes, and propagating uncertainty into spatial predictions. To our knowledge, no previous study in Malawi has applied MBG to continuous haemoglobin measurements to map fine-scale anaemia risk in direct relation to individual-level S. haematobium infection intensity. Chipeta et al. [12] applied spatial modelling to S. haematobium infection data from 18 villages in Chikhwawa district, southern Malawi, mapping variation in infection prevalence and intensity among 1,642 participants but did not incorporate haemoglobin measurements or examine anaemia as an outcome. Similarly, Seiler et al.[13] provided high-resolution spatio-temporal estimates of childhood anaemia across 37 low- and middle-income countries using Bayesian distributional regression but did not specifically model S. haematobium infection intensity as a driver of anaemia, nor examine village-level spatial heterogeneity in Malawi. Our study therefore addresses a critical gap by integrating these two dimensions at fine village-level spatial resolution.

This spatial mapping gap is compounded by a further methodological limitation. Previous anaemia mapping studies have routinely dichotomised haemoglobin levels, even though predicting anaemia prevalence does not require converting Hb into a binary outcome. The use of a dichotomous indicator for anaemia obtained from thresholding Hb measurements discards substantial information. Some studies have suggested that dichotomization leads to losses of up to 36% of Fisher’s information which in turn can reduce the precision of regression estimates [14, 15]. As a result, spatial predictions for anaemia can also be negatively impacted, masking subtle geographic gradients in risk and limiting our ability to capture fine-scale spatial patterns of anaemia burden[14–16].

This study addresses these critical limitations. We utilise linear Model-Based Geostatistics (MBG) applied directly to continuous haemoglobin concentrations from the HUGS investigation to retain maximum statistical power, thereby overcoming the imprecision inherent in dichotomised models. Critically, this is the first study in Malawi to integrate this rigorous spatial approach with detailed, individual-level *S. haematobium* infection intensity data (eggs/10 mL categories). Our objectives were two-fold: (1) to quantitatively determine the fine-scale association between urogenital schistosomiasis intensity and severity of anaemia and (2) to generate high-resolution spatial maps and exceedance probabilities that identify precise, high-risk clusters to guide future targeted interventions, including optimisation of preventive chemotherapy delivery and complementary public health strategies.

## Methods

### Ethics statement

The study protocol, including the informed consent forms, was reviewed and approved by the Kamuzu University of Health Sciences (KUHeS) Research Ethics Committee (KUREC) (Approval # P.08/21/3381) and the Liverpool School of Tropical Medicine (LSTM) Research Ethics Committee prior to commencement of the study. The present analysis used anonymised data collected under these approvals.

### Study design and population

The study used data from the HUGS investigation that explored the evolving epidemiology of schistosomiasis in two sentinel village communities: Samama and Mthawira, located in Mangochi and Nsanje districts, respectively. Data were collected as part of the first annual community survey conducted in 2022, with participants eligible if they were aged 2 years and above and permanent residents of the study villages. Full details of the HUGS study design and data collection procedures are available here [9–11].

For this analysis, baseline community-based data were collected from locations selected based on schistosomiasis risk, and to reflect variation in age and sex. Approximately 1,200 participants were invited from each district: the areas surrounding Samama School in Mangochi and Mthawira School in Nsanje, of whom 1,149 from Samama and 977 from Mthawira provided complete data for this analysis. These sites were selected due to the high prevalence of urogenital schistosomiasis and prior discovery of hybrid *Schistosoma* species.

### Specimen collection, laboratory analysis

Capillary blood was collected via finger prick using sterile lancets. Hb concentration (g/L) was measured on-site using the HemoCue® Hb 801 system. Participants identified as anaemic were tested for malaria using a rapid diagnostic test (mRDT). Those testing positive were treated on-site and excluded from the analysis to avoid malaria-related confounding.

Urine specimens were collected in sterile containers labeled with anonymous IDs and processed in a temporary field laboratory. For detection of eggs, 10 mL of urine was filtered through polycarbonate membrane filters using sterile syringes. The filtrate was examined under light microscopy at 10× and 40× magnification. Results were categorised as negative; light (0–9 eggs); moderate (10–49 eggs); or heavy (≥50 eggs) per 10 mL of urine. Any incidental findings of *S. mansoni* or atypical eggs were also recorded. All data were securely collected and uploaded using Kobo Toolbox (version 2022.2.3) and stored in a centralised electronic database.

### Individual and environmental-level predictors

Individual- and household-level data were collected using a structured questionnaire administered on electronic tablets (Samsung Electronics Co., Ltd.) and programmed in Kobo Toolbox (version 2022.2.3). The questionnaire captured participants’ demographic and socio-economic characteristics, as well as information on knowledge, attitudes, and practices related to WHO-recommended schistosomiasis prevention and control interventions. For the purposes of this analysis, individual-level covariates included age, sex, household size, included as an indicator of household crowding and a proxy for resource constraints, consistent with its application in similar sub-Saharan African epidemiological studies[4, 17], and S. haematobium infection intensity.

Spatial covariates included population density, elevation, and distance to water bodies. Population density and elevation data were obtained from WorldPop (2020)[18], using ∼100 m resolution raster layers matched to participant GPS coordinates. Distance to water bodies, defined as the Euclidean distance from each household to the nearest river, lake, or permanent water source, was derived from shapefiles obtained from ArcGIS[19]. All variables were selected based on biologically plausible associations with urogenital schistosomiasis transmission and anaemia prevalence, consistent with existing literature[5, 17, 20, 21].

### Geostatistical modeling

To quantify the association between haemoglobin levels and both individual and environmental risk factors, while accounting for spatial variation, we applied a Gaussian geostatistical model following the framework described by Kyomuhangi et al. [15]. Unlike conventional approaches that dichotomise haemoglobin into anaemic versus non-anaemic categories, our model retains haemoglobin as a continuous outcome, thereby preserving statistical power and capturing more nuanced relationships.

We modeled the natural logarithm of haemoglobin concentration, log (Hb₍ᵢⱼ₎), for individual *j* at location *xᵢ*, as a linear function of age, gender, household size, infection intensity, population density, elevation, and distance to water bodies as described in below equation.

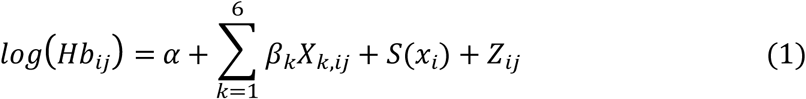

Here, the intercept is represented by *α*; *X_k_*_,*ij*_ represents the *k*-th covariate for individual *j* at location *i*, with associated coefficient *β_k_*; *S*(*x_i_*) is a stationary, isotropic Gaussian process capturing unmeasured spatially structured variation for location *x_i_*; and *Z_ij_* is an independent random error term capturing individual-level variation. The Gaussian process *S*(*x_i_*) was modelled with an exponential covariance function, parameterised by variance *σ*^2^ and spatial scale φ, which controls the rate of spatial correlation decay with distance. Model parameters were estimated using maximum likelihood estimation (MLE), implemented in R statistical software[22] using the RiskMap package (version 1.0.0)[23, 24], which implements stationary Gaussian processes with Matérn correlation for spatial prediction. Critically, anaemia prevalence at each location was derived directly from the continuous haemoglobin model without requiring prior dichotomisation of individual measurements. Following the derivation in S1 Text, the predicted probability that an individual’s haemoglobin falls below the WHO age- and sex-specific threshold c at location xᵢ was obtained by incorporating the log-transformed threshold as an additional covariate in the linear predictor. This approach transforms the continuous model into a binomial geostatistical model with a probit link function (S1 Text), directly propagating uncertainty from the continuous haemoglobin predictions into prevalence estimates, thereby addressing the statistical limitations of conventional dichotomisation [13–15]. Full model specification, assumptions, and derivations for the binary anaemia model transformation and threshold application are provided in **S1 Text**.

### Spatial predictions for prevalence of anaemia among girls aged 6–12 years

To predict the risk of anaemia in girls aged 6–12 years, we applied the WHO haemoglobin threshold of <110 g/L, which accounts for individual characteristics such as age, sex, and pregnancy status (**S1 Table**). Exceedance probability maps were produced representing the posterior probability that anaemia prevalence exceeded a pre-specified threshold at each location, 20% for Mangochi, consistent with the WHO criterion for anaemia of moderate public health significance[25], and 50% for Nsanje, where the elevated baseline prevalence necessitated a higher threshold to reveal meaningful spatial variation. This age group was selected because moderate anaemia prevalence was substantially higher among females aged 6–12 years compared with other age-sex subgroups, exceeding 45% in Samama and 39% in Mthawira, making them a particularly vulnerable subgroup for targeted interventions.

## Results

A total of 2,280 participants provided urine samples for S. haematobium ova examination, whilst 2,421 provided blood samples for haemoglobin (Hb) analysis. After accounting for incomplete data and excluding participants who tested positive for malaria, 1,149 participants from Mangochi and 977 from Nsanje were included in the final analysis. Only participants with complete geographical (location coordinates), demographic (age and gender), and morbidity (haemoglobin concentration and schistosomiasis infection intensity) information were included. **Fig 1** illustrates the study districts within Malawi and the distribution of participants across the sampled households.

**Fig 1.**
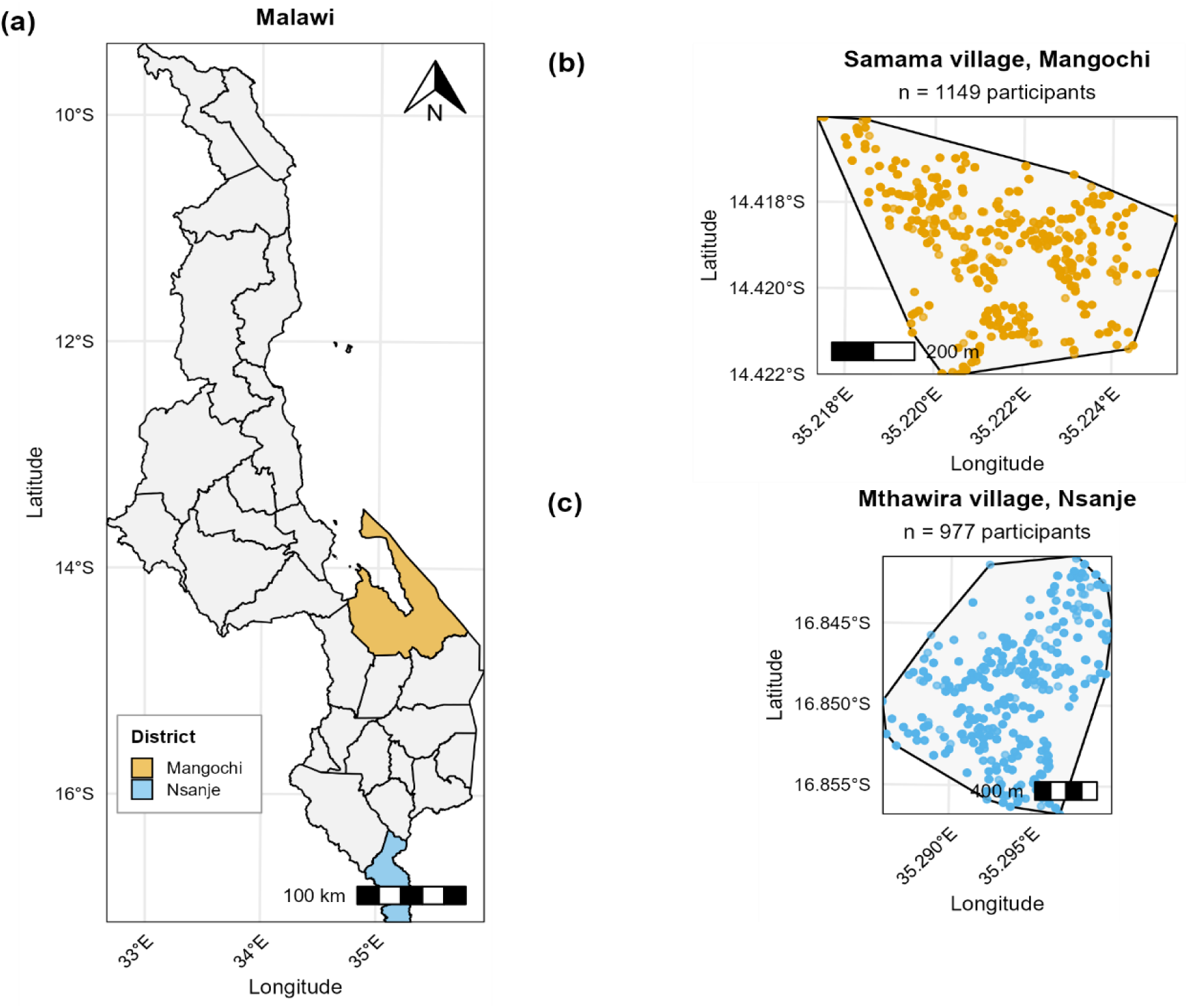
Location of study sites and spatial distribution of survey participants in Malawi. Panel (a) shows the geographical location of the two study districts within Malawi: Mangochi district (orange), situated at the southern end of Lake Malawi, and Nsanje district (blue), located within the Lower Shire River Valley. Panels (b) and (c) show the spatial distribution of surveyed participants by household location in Samama village, Mangochi district (n = 1,149) and Mthawira village, Nsanje district (n = 977), respectively. Each dot represents the GPS-recorded household location of one participant. Polygon boundaries delineate the convex hull of the surveyed area within each village. Scale bars indicate distances in metres.

The Mean age of the participants was 15 years (SD ± 11), and the median household size was 5 members (IQR: 4–6). The spatial distribution of haemoglobin (Hb) concentrations across 1,149 and 977 locations in Samama and Mthawira, respectively, is presented in **Fig 2**.

**Fig 2.**
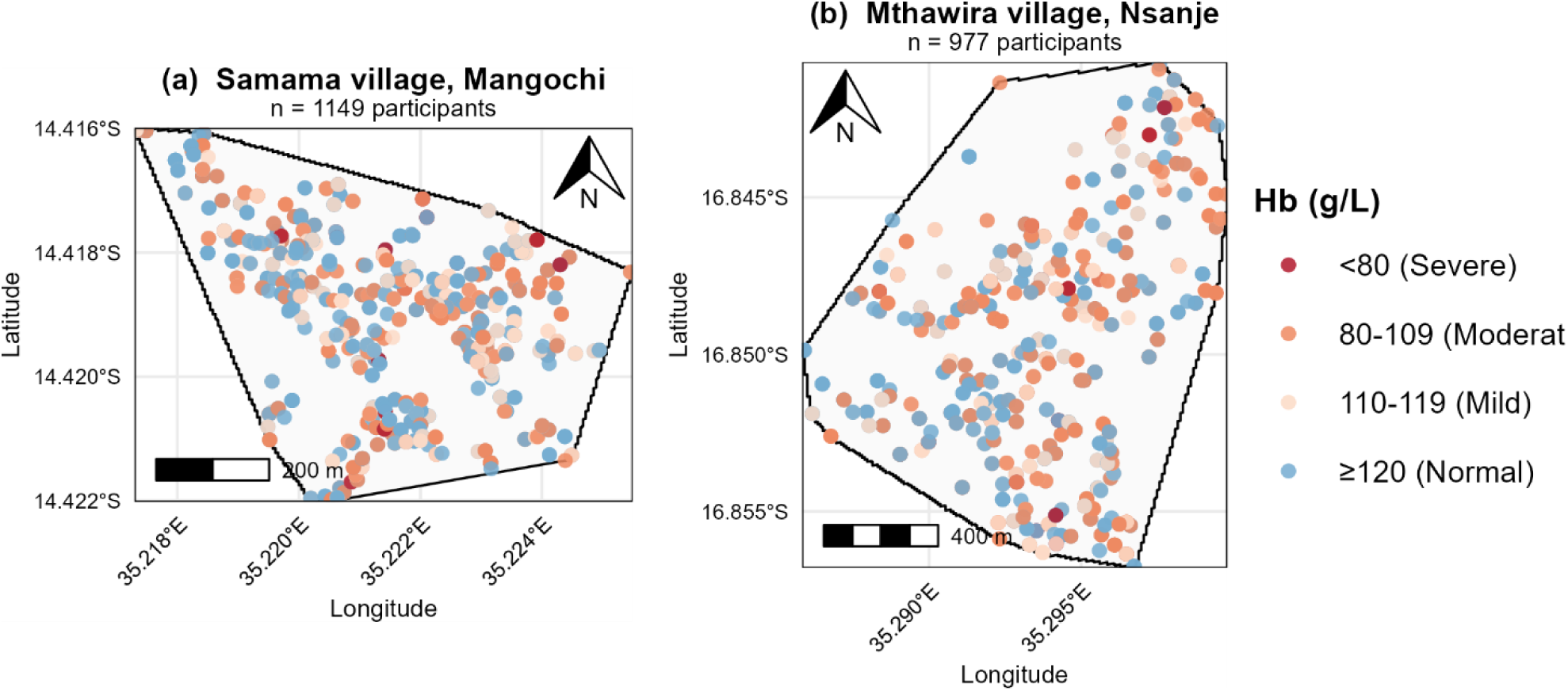
Spatial distribution of haemoglobin (Hb) concentrations by household location in Samama village, Mangochi (a) and Mthawira village, Nsanje (b). Each dot represents the GPS-recorded household location of one participant, coloured according to WHO anaemia severity classification: dark red (<80 g/L, severe anaemia), orange (80–109 g/L, moderate anaemia), light peach (110–119 g/L, mild anaemia), and light blue (≥120 g/L, normal Hb). Polygon boundaries delineate the convex hull of the surveyed study area within each village. Scale bars indicate distances in metres. Hb = haemoglobin; g/L = grams per litre.

The prevalence of mild, moderate, and severe anaemia in Samama was 20.1%, 20.3%, and 1.6%, respectively, whereas in Mthawira, the prevalence was slightly higher, with 23.2% having mild anaemia, 27.0% with moderate anaemia, and 1.9% with severe anaemia. Analysis of Haemoglobin levels by age revealed that Hb levels increased gradually with age across both sites (**Fig 3**).

**Fig 3:**
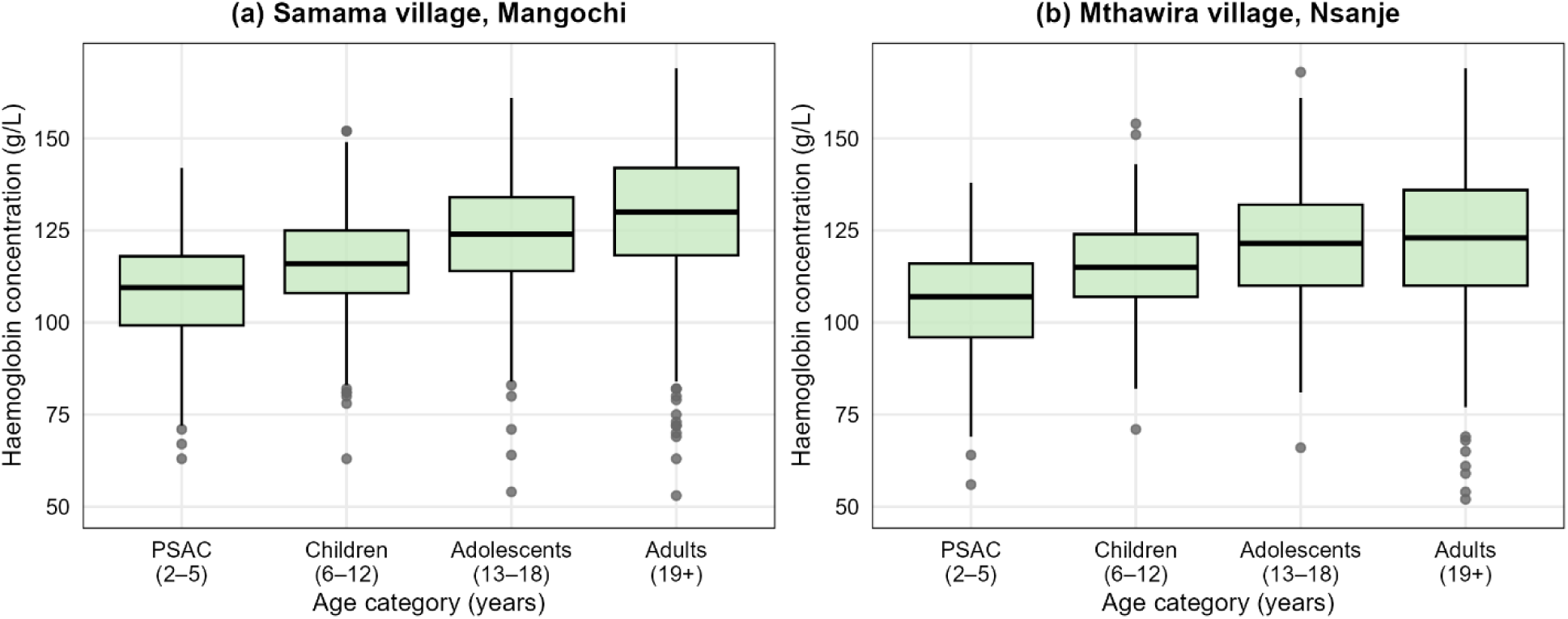
Haemoglobin concentrations by age category in Samama village, Mangochi (a) and Mthawira village, Nsanje (b). Boxplots display the distribution of haemoglobin (Hb) concentrations (g/L) across four age categories, pre-school-aged children (PSAC, 2–5 years), children (6–12 years), adolescents (13–18 years), and adults (≥19 years). The horizontal line within each box represents the median Hb concentration, box edges represent the interquartile range (IQR), whiskers extend to 1.5 × IQR beyond the box edges, and individual dots represent outliers. Hb = haemoglobin; g/L = grams per litre; PSAC = pre-school-aged children; IQR = interquartile range.

The highest prevalence of moderate anaemia was observed among children aged 6–12 years, accounting for 45% in Samama and 40% in Mthawira. In contrast, mild and severe anaemia were more prevalent among adults aged 19 years and older, a trend consistent across both study sites. Gender-based distribution indicated that the proportion of females with anaemia (mild, moderate, and severe) was comparatively higher than males in both sites. The detailed descriptives of study participants were presented in **Table 1a** and **Table 1b**.

**Table 1.** Sociodemographic and clinical characteristics of study participants by anaemia severity in Samama village, Mangochi district.

| Characteristics | Overall<br>N = 1,149 | No anaemia<br>666 (58.0%) | Mild<br>231 (20.1%) | Moderate<br>234 (20.3%) | Severe<br>18 (1.6%) |
| --- | --- | --- | --- | --- | --- |
| **Age (years)** |  |  |  |  |  |
| Mean (SD) | 15 (11) | 16 (11) | 15 (11) | 11 (9) | 21 (14) |
| **Age category** |  |  |  |  |  |
| PSAC (2–5 years) | 214 (19.0%) | 101 (15.2%) | 51 (22.1%) | 58 (25.0%) | 4 (22.2%) |
| Children (6–12 years) | 368 (32.1%) | 206 (31.0%) | 54 (23.0%) | 106 (45.2%) | 2 (11.1%) |
| Adolescents (13–18 years) | 238 (21.0%) | 128 (18.8%) | 74 (32.0%) | 33 (14.1%) | 3 (17.2%) |
| Adults (≥19 years) | 329 (28.9%) | 231 (35.0%) | 52 (23.0%) | 37 (15.7%) | 9 (49.5%) |
| **Sex** |  |  |  |  |  |
| Male | 509 (44.6%) | 275 (41.3%) | 123 (53.2%) | 107 (46.0%) | 4 (22%) |
| Female | 640 (56.4%) | 391 (58.6%) | 108 (46.8%) | 127 (54.0%) | 14 (78%) |
| **Household size** |  |  |  |  |  |
| Median (Q1, Q3) | 5 (4, 6) | 5 (4, 6) | 5 (4, 6) | 5 (4, 6) | 4.5 (3, 6) |
| **S. haematobium infection intensity** |  |  |  |  |  |
| Negative | 553 (47.8%) | 340 (50.7%) | 100 (43.3%) | 106 (44.9%) | 7 (38.5%) |
| Light (0–9 eggs/10 mL) | 266 (23.2%) | 163 (24.1%) | 54 (23.4%) | 45 (19.0%) | 4 (22.3%) |
| Moderate (10–49 eggs/10 mL) | 188 (16.0%) | 90 (14.1%) | 45 (19.5%) | 49 (21.1%) | 4 (22.2%) |
| Heavy (≥50 eggs/10 mL) | 142 (12.0%) | 73 (11.0%) | 32 (13.9%) | 34 (15.0%) | 3 (17.0%) |

Data presented as n (%) unless otherwise stated. Anaemia classified according to WHO haemoglobin thresholds for the relevant age and sex group: mild anaemia (Hb 110–119 g/L for most groups), moderate anaemia (Hb 80–109 g/L), and severe anaemia (Hb <80 g/L).
Abbreviations: PSAC, pre-school-aged children; SD, standard deviation; Q1, first quartile; Q3, third quartile; Hb, haemoglobin; g/L, grams per litre.

**Table 2.**
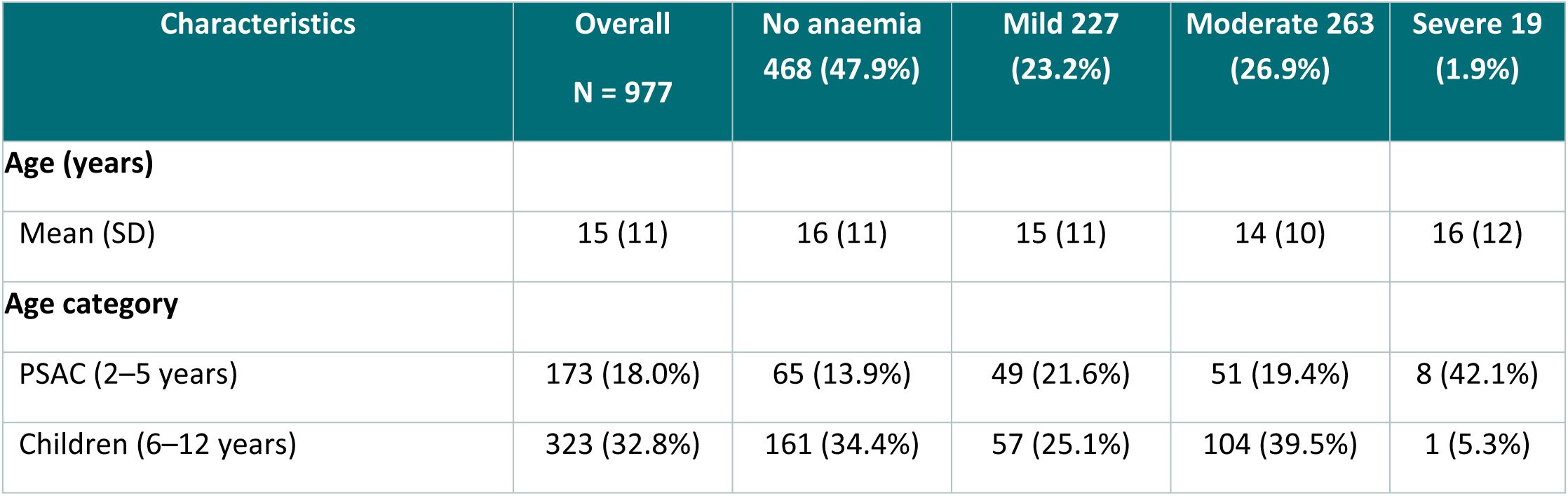
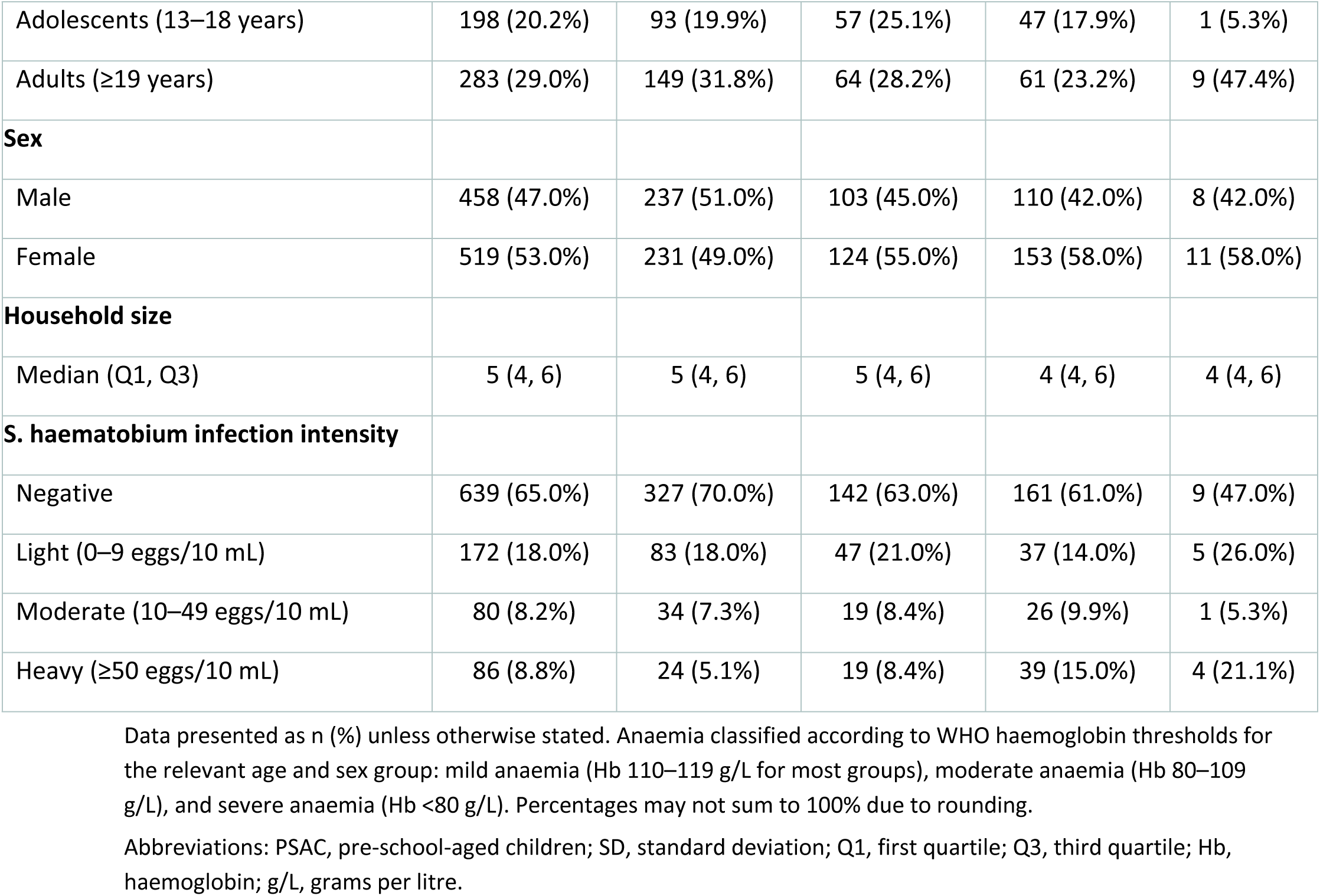
Sociodemographic and clinical characteristics of study participants by anaemia severity in Mthawira village, Nsanje district.

Urine microscopy showed differences in schistosomiasis infection intensity between the two sites and its association with anaemia severity. Participants with moderate anaemia had a higher prevalence of 10–49 eggs per 10 ml of urine, accounting for 21% in Samama and 10% in Mthawira. Conversely, those with severe anaemia exhibited a higher proportion of ≥50 eggs per 10 ml of urine, with 17% in Samama and 21% in Mthawira. The relationship between schistosomiasis infection intensity and Haemoglobin concentration is illustrated in **Fig 4**.

**Fig 4:**
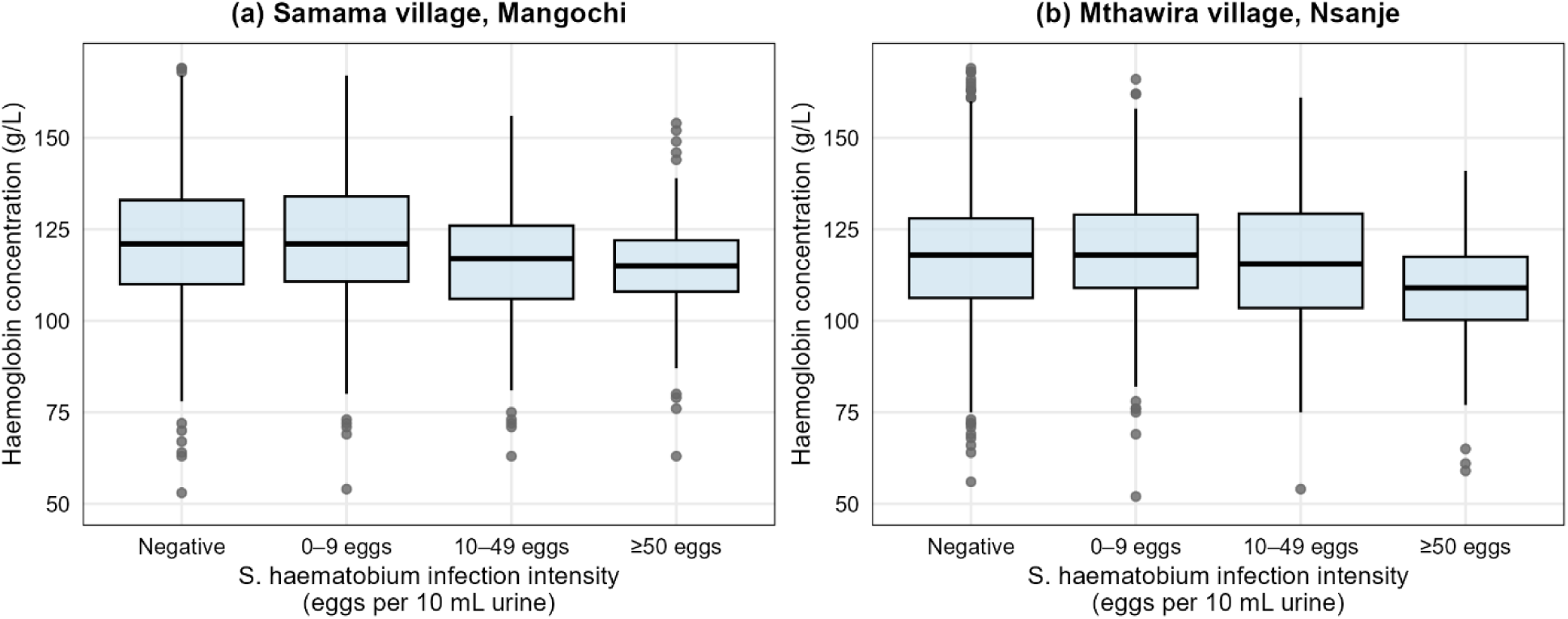
Distribution of haemoglobin concentrations by *S. haematobium* infection intensity in Samama village, Mangochi (a) and Mthawira village, Nsanje (b). Boxplots display haemoglobin (Hb) concentrations (g/L) across four infection intensity categories, negative, light (0–9 eggs/10 mL), moderate (10–49 eggs/10 mL), and heavy (≥50 eggs/10 mL), based on urine microscopy. The horizontal line within each box represents the median, box edges represent the interquartile range (IQR), whiskers extend to 1.5 × IQR, and dots represent outliers. Hb = haemoglobin; g/L = grams per litre.

### Geostatistical modeling

Maximum likelihood estimates from the linear geostatistical model are presented in **Table 3**. Estimates were exponentiated to the original scale for interpretation. Model estimates confirmed descriptive patterns, showing a strong positive association between age and haemoglobin (Hb) across both districts (all age groups p < 0.001 vs 2–5 years). Females had significantly lower Hb than males in both Mangochi (β = 0.95, 95% CI: 0.95–0.96) and Nsanje (β = 0.96, 95% CI: 0.96–0.96). Household size was not significant in Mangochi but showed a small positive effect in Nsanje (β = 1.01, 95% CI: 1.01–1.02, p = 0.008).

**Table 2.** Maximum likelihood estimates from geostatistical model for Mangochi and Nsanje.

| Covariate | Mangochi Estimate (95% CI) | Mangochi p-value | Nsanje Estimate (95% CI) | Nsanje p-value |
| --- | --- | --- | --- | --- |
| Intercept | 128.42 (120.08, 137.34) | <0.001 | 89.73 (47.57, 169.25) | <0.001 |
| **Age category (ref: PSAC 2–5 years)** |  |  |  |  |
| Children (6–12 years) | 1.08 (1.07, 1.10) | <0.001 | 1.03 (1.02, 1.04) | <0.001 |
| Adolescents (13–18 years) | 1.15 (1.14, 1.16) | <0.001 | 1.05 (1.05, 1.06) | <0.001 |
| Adults ($\geq 19$ years) | 1.20 (1.19, 1.21) | <0.001 | 1.08 (1.07, 1.08) | <0.001 |
| **Sex (ref: Male)** |  |  |  |  |
| Female | 0.95 (0.95, 0.96) | <0.001 | 0.958 (0.956, 0.961) | <0.001 |
| Household size | 1.00 (0.99, 1.01) | 0.435 | 1.01 (1.01, 1.02) | 0.008 |
| **Infection intensity (ref: Negative)** |  |  |  |  |
| Light (0–9 eggs/10 mL) | 1.006 (1.005, 1.009) | <0.001 | 0.984 (0.982, 0.987) | <0.001 |
| Moderate (10–49 eggs/10 mL) | 0.971 (0.968, 0.974) | <0.001 | 0.975 (0.972, 0.978) | <0.001 |
| Heavy ($\geq 50$ eggs/10 mL) | 0.988 (0.985, 0.992) | <0.001 | 0.952 (0.949, 0.954) | <0.001 |
| **Environmental covariates** |  |  |  |  |
| Elevation (m) | 0.0026 (0.0033, 0.018) | <0.001 | 0.0083 (0.0073, 0.0094) | <0.001 |
| Population density | 1.016 (1.015, 1.017) | <0.001 | 1.007 (1.007, 1.008) | <0.001 |
| Distance to nearest water body (m) | 0.991 (0.991, 1.010) | <0.001 | 1.00 (1.00, 1.01) | <0.001 |
| **Spatial parameters** |  |  |  |  |
| Nugget variance | 0.0196 (0.0195, 0.0198) | — | 0.0101 (0.0101, 0.0102) | — |
| Spatial scale $\phi$ (km) | 25.5 (24.4, 27.4) | — | 66.7 (44.5, 99.8) | — |
| Spatial variance $\sigma^2$ | $5.0 \times 10^{-4}$<br>($5.1 \times 10^{-4}$, $6.0 \times 10^{-4}$) | — | $5.0 \times 10^{-5}$<br>($3.0 \times 10^{-5}$, $1.0 \times 10^{-4}$) | — |

Estimates represent exponentiated coefficients from the linear geostatistical model applied to log-transformed haemoglobin concentrations. Values greater than 1.0 indicate higher haemoglobin relative to the reference category; values less than 1.0 indicate lower haemoglobin. Spatial parameters are not subject to hypothesis testing and are therefore presented without p-values (—).
Abbreviations: CI, confidence interval; PSAC, pre-school-aged children; ref, reference category; $\phi$ , spatial scale parameter (km); $\sigma^2$ , spatial variance; m, metres; Hb, haemoglobin.

Infection intensity patterns differed by site. In Mangochi, low-intensity infection (0–9 eggs) was associated with slightly higher Hb (β = 1.006, 95% CI: 1.005–1.009), while moderate (10–49 eggs, β = 0.971, 95% CI: 0.968–0.974) and heavy infections (≥50 eggs, β = 0.988, 95% CI: 0.985–0.992) were associated with reduced Hb. In Nsanje, all infection intensities were linked to lower Hb: 0– 9 eggs (β = 0.984), 10–49 eggs (β = 0.975), and ≥50 eggs (β = 0.952), all p < 0.001. Among spatial covariates, population density was positively associated with Hb in both districts. Elevation also showed a small but positive effect. In contrast, distance to the nearest water body was negatively associated with Hb in Mangochi and showed no clear association in Nsanje. The estimated spatial variance was higher in Mangochi (σ² = 5.0×10⁻⁴) than Nsanje (σ² = 5.0×10⁻⁵), while spatial correlation extended further in Nsanje (φ = 66.6 km) compared to Mangochi (φ = 25.5 km).

### Predicted risk of anaemia among female children aged 6–12 years

Spatial predictions of anaemia prevalence among female children aged 6–12 years with moderate *S. haematobium* infection intensity (10–49 eggs/10 mL) and from five-member households reveal distinct patterns across Mangochi and Nsanje (**Fig 5a and 5b**). In Mangochi, predicted anaemia prevalence was moderate to high, ranging from approximately 25% to 65%, with higher prevalence concentrated in the central and north-western areas of the village. The corresponding exceedance probability map indicates spatially heterogeneous clusters where the probability that anaemia prevalence exceeds 20% is greater than 75%, highlighting localised high-risk areas within the village that are priorities for targeted intervention.

**Fig 5.**
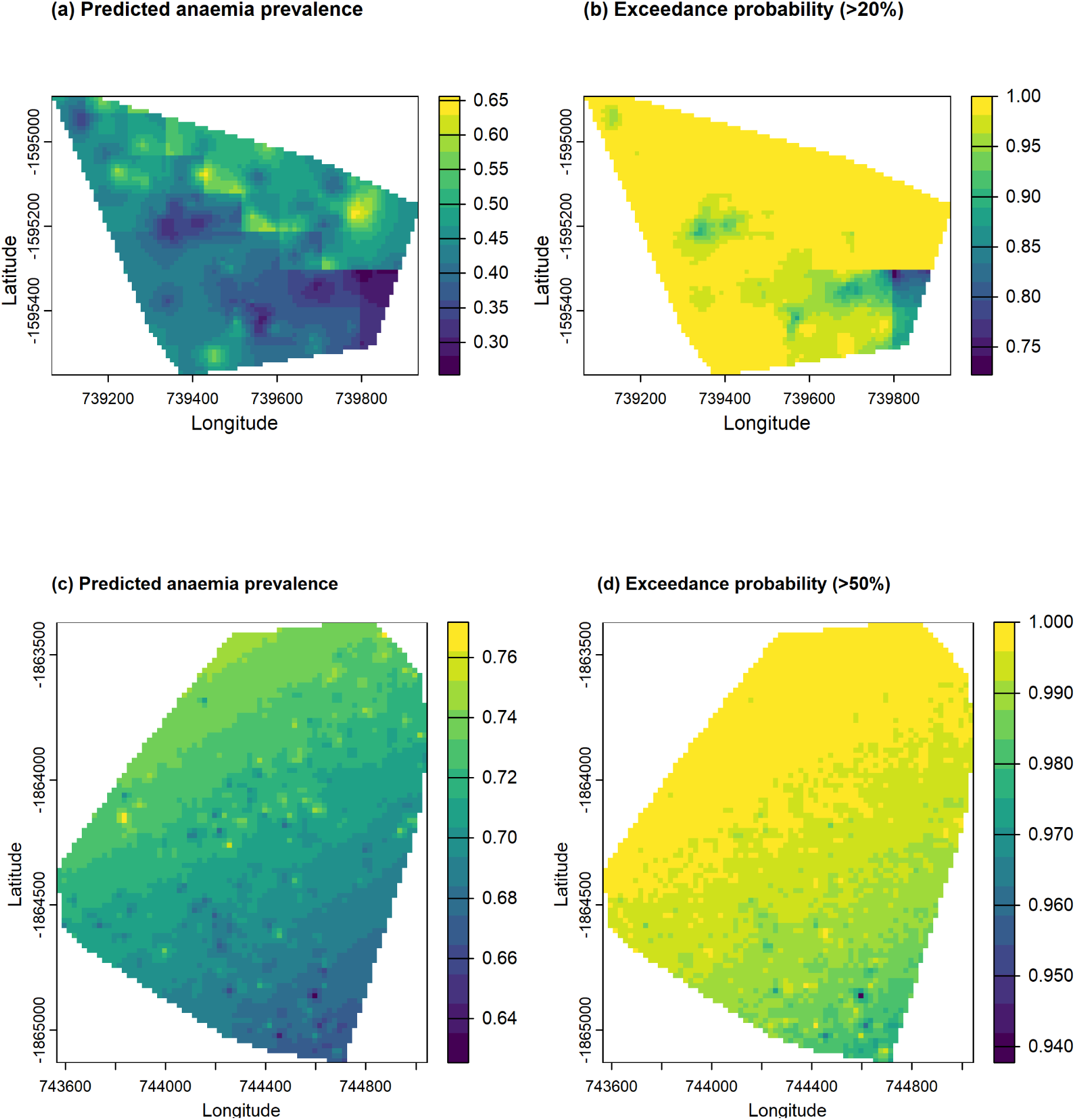
Predicted anaemia prevalence and exceedance probability maps for female children aged 6–12 years with moderate *S. haematobium* infection intensity in Samama village, Mangochi (a, b) and Mthawira village, Nsanje (c, d). Panels (a) and (c) show the posterior mean predicted anaemia prevalence, defined as haemoglobin concentration below 110 g/L — across each study area, derived from the linear geostatistical model applied to continuous haemoglobin measurements. Panels (b) and (d) show the exceedance probability maps, representing the posterior probability that anaemia prevalence exceeds 20% at each location in Mangochi and 50% in Nsanje respectively. The higher threshold was applied in Nsanje to account for the substantially elevated baseline prevalence in that community, where a 20% threshold yielded near-universal exceedance with no meaningful spatial discrimination. High exceedance probabilities identify priority areas for targeted intervention. Colour ranges from low (dark purple) to high (yellow). Coordinates are metres (UTM Zone 36S). Hb = haemoglobin; g/L = grams per litre.

In Nsanje, predicted anaemia prevalence was substantially higher overall, ranging from approximately 64% to 76%, reflecting the greater burden of anaemia in this setting. Higher prevalence was observed primarily in the northern and central areas of the village. Given the elevated baseline prevalence, a higher exceedance threshold of 50% was applied; exceedance probability maps show consistently high probabilities (>94%) that anaemia prevalence exceeds 50% across much of the site, reflecting spatially persistent and uniformly high risk with relatively low prediction uncertainty. Overall, Nsanje exhibits a more severe and spatially extensive anaemia burden compared with Mangochi, indicating a district-wide public health emergency requiring immediate and sustained intervention.

## Discussion

With the increasing burden of anaemia in sub-Saharan Africa (SSA), there is a pressing need to enhance the understanding of how to monitor and manage efficiently the multiple causes of anaemia, particularly in schistosomiasis-endemic regions. This study examined the association between anaemia severity and *S. haematobium* infection intensity among 2,126 participants across two sites, Samama and Mthawira, located in the Mangochi and Nsanje districts of Malawi.

We employed a linear geostatistical model without dichotomising Haemoglobin levels into binary outcomes, allowing us to quantify the risk of anaemia by defining a continuous Haemoglobin threshold below 110 g/L. This approach is particularly useful when anaemia cut-offs vary across individuals due to factors such as age, sex, and pregnancy status, which are often unaccounted for in binary models used in previous anaemia mapping studies [14, 15]. Ignoring such variability can lead to inaccurate classification and biased estimates, especially when critical covariates such as pregnancy status are missing. Additionally, the anaemia thresholds used in clinical practice, as outlined by the World Health Organization (WHO) guidelines from 1992-2001, may be subject to revision as scientific evidence evolves, further highlighting the importance of employing flexible, data-driven models[15, 16].

The findings from this analysis highlighted key differences in anaemia prevalence, infection intensity, and geographical variations, providing crucial insights into the burden of anaemia and schistosomiasis intensity in these regions. The prevalence of mild, moderate, and severe anaemia was found to be higher in Mthawira compared to Samama, with 27.0% of participants in Mthawira exhibiting moderate anaemia versus 20.3% in Samama. The highest burden of moderate anaemia was observed among children aged 6–12 years, accounting for 45% in Samama and 40% in Mthawira. This pattern is consistent with previous studies that have reported a higher susceptibility to anaemia in school-aged children due to nutritional deficiencies and parasitic infections, particularly schistosomiasis[3, 20, 26]. The higher infection burden in this age group may be attributed to increased exposure to contaminated water sources and limited access to preventive measures, such as mass drug administration and health education [8]. Gender-based analysis revealed a higher prevalence of anaemia among females in both Samama and Mthawira, which aligns with global trends where women, particularly of reproductive age, are more vulnerable to anaemia due to menstrual blood loss and increased iron demands during pregnancy [26].

Urine microscopy revealed a strong association between *S. haematobium* infection intensity and anaemia severity, with participants exhibiting higher egg counts (≥50 eggs/10 mL) showing a significant reduction in Hb levels (p < 0.001). This highlights the detrimental impact of urogenital schistosomiasis on hematological health, particularly in contributing to anaemia. The higher prevalence of moderate anaemia among participants with 10–49 eggs/10 mL and severe anaemia among those with ≥ 50 eggs/10 mL suggests that the physiological burden imposed by *S. haematobium* directly contributes to reduced Hb concentrations. This pattern is consistent with previous research demonstrating that schistosomiasis can lead to chronic blood loss, nutritional deficiencies, and immune-mediated destruction of erythrocytes, culminating in anaemia[27, 28]. The inclusion of *S. haematobium* infection intensity as a significant covariate in the geostatistical model further reinforces the potential utility of urine egg count as a predictor for mapping anaemia severity, particularly in endemic regions such as Malawi [3].

Geographically, the relationship between schistosomiasis infection and anaemia was more spatially variable in Mangochi, reflecting localised heterogeneity likely driven by fine-scale differences in water contact patterns and household-level risk factors. Spatial analysis revealed significant variability in anaemia across the two districts, with stronger and longer-range spatial correlation observed in Nsanje (φ = 66.7 km, 95% CI: 44.5–99.8) compared with Mangochi (φ = 25.5 km, 95% CI: 24.4–27.4), indicating more spatially persistent risk in Nsanje and more localised heterogeneity in Mangochi. The estimated model was used to predict the prevalence of anaemia among female children aged 6–12 years with moderate infection intensity (10–49 eggs/10 mL) from five-member households. In Mangochi, the predicted prevalence ranged from approximately 25% to 65%, with high-risk clusters concentrated in the central and north-western areas of the village. In Nsanje, predicted prevalence was substantially higher overall, ranging from 64% to 76%, reflecting the greater anaemia burden in this setting. Exceedance probability maps revealed near-uniform high probabilities exceeding 94% across most of the Nsanje study area when applying a 50% threshold, while in Mangochi, exceedance probabilities for the 20% threshold showed meaningful spatial heterogeneity, with high-risk clusters in the northern and central areas. These findings highlight the heterogeneity of schistosomiasis-associated anaemia risk in Malawi, likely reflecting differences in environmental conditions, water contact patterns, and access to healthcare across the two settings[17].

Interestingly, population density was positively associated with the risk of anaemia in both Mangochi and Nsanje (p < 0.001), suggesting that participants in densely populated areas may have better access to healthcare services, nutritional resources, and community-based interventions, which may mitigate some of the risk factors associated with anaemia. This finding aligns with previous studies that have demonstrated a positive association between population density and healthcare access, as individuals in urban or densely populated areas are more likely to have better socioeconomic status and healthcare utilization[17, 29]. Similarly, distance to water bodies was found to be inversely associated with the risk of anaemia, indicating that participants residing closer to water bodies had a higher likelihood of anaemia [7]. This relationship is likely attributable to increased exposure to *S. haematobium* through frequent water contact, particularly in communities where water bodies serve as primary sources for domestic activities such as bathing, washing, and fetching water[8].

This study leveraged detailed geospatial data, robust statistical modeling, and a large sample size to assess the relationship between anaemia, schistosomiasis, and demographic factors. Given the high prevalence of anaemia among school-aged children and females, school-based deworming programs, iron supplementation, and improved access to clean water and sanitation should be prioritized. The identification of high-risk spatial clusters through exceedance probability mapping highlights the importance of usage of geostatistical models and geographically targeted interventions, which can maximize resource efficiency and improve health outcomes.

Despite the valuable contributions of this study, several limitations should be acknowledged. First, the cross-sectional design prevents establishing causality, and residual confounding from unmeasured factors such as nutritional status, socioeconomic status, and co-infections cannot be ruled out. Second, despite excluding malaria RDT-positive participants, sub-patent malaria infections may have contributed to anaemia and could not be fully excluded as a confounder. Third, pregnancy status was not fully ascertained in all female participants, which may have affected haemoglobin threshold classification in women of reproductive age. Fourth, while urine filtration microscopy was appropriate for quantifying S. haematobium egg burden in a field setting, its sensitivity is reduced in low-intensity infections where egg excretion may be intermittent, potentially leading to underestimation of true infection prevalence; a single urine sample was collected per participant, whereas duplicate samples are recommended by WHO to improve diagnostic accuracy. The findings from these two sentinel communities in southern Malawi may not be directly generalisable to all schistosomiasis-endemic settings given the distinct ecological and epidemiological characteristics of each site. However, the geostatistical framework applied here is transferable to other high-burden settings and could inform national-level anaemia mapping programmes in Malawi and similar endemic countries.

## Conclusion

The study provides compelling evidence of the spatial heterogeneity of anaemia risk and its strong association with urogenital schistosomiasis infection intensity across Mangochi and Nsanje. By identifying high-risk clusters and vulnerable populations, this research highlights the need for targeted public health interventions to mitigate anaemia and its associated morbidities in these endemic regions. Additionally, the use of geostatistical modeling offers a valuable data-driven tool to predict anaemia risk, enabling more effective allocation of resources and intervention strategies.

## Data Availability

The minimal dataset and R code underlying the findings of this study will be made available in a public repository upon acceptance. Data requests can be made to the corresponding author at prior to that

## Acknowledgement

The authors would like to acknowledge support for this study that was received from the following governmental bodies and persons: District Commissioners, Directors of Health and Social Services, District Medical Officers, District Environmental Health Officers, District Schistosomiasis Coordinators, Management and staff of Mangochi and Nsanje District Councils; the Assistant Environmental Health Officers, Health Centre officers in charge and Health Surveillance Assistants of Mpondasi and Tengani Health Centres. Additionally, we would like to extend special thanks to the Headteachers and teachers of Samama and Mthawira Primary Schools, Research Assistants, all community leaders and members of the two communities who participated in the study. Also, we thank all core members of the HUGS team in Malawi, and especially Mr Andrew Nguluwe, of the NSCP, for the unwavering support rendered to the study.

## Author contributions

FK: Conceptualisation, Data curation, Formal analysis, Methodology, Software, Visualisation, Writing original draft, Writing, review and editing. SAK: Data curation, Investigation, Project administration, Writing, review and editing. PM: Data curation, Investigation, Writing, review and editing. JM: Conceptualisation, Funding acquisition, Project administration, Resources, Supervision, Writing, review and editing. SR: Data curation, Investigation, Writing, review and editing. JRS: Conceptualisation, Funding acquisition, Resources, Supervision, Writing, review and editing. AB: Data curation, Investigation, Writing, review and editing. EG: Conceptualisation, Methodology, Supervision, Writing, review and editing.

## Notes

### Competing Interest Statement

The authors have declared no competing interest.

### Author Declarations

This study was approved by the Kamuzu University of Health Sciences Research Ethics Committee (KUHeS), Malawi (approval number: P.08/21/3381) and the Liverpool School of Tropical Medicine Research Ethics Committee (LSTM REC), United Kingdom. Written informed consent was obtained from all participants prior to enrolment. For participants aged below 18 years, written informed consent was obtained from a parent or guardian

